# Mortality and predictors among children aged 2 to 59 months admitted with severe pneumonia at the Regional Referral Hospital of Banfora, Burkina Faso

**DOI:** 10.1101/2025.11.17.25340456

**Authors:** Kiebré Pegd-Wendé Blaise, Dahourou Desiré Lucien, Bountogo Mamadou, Dah Ter Tiero Elias, Tall Haoua, Ouedraogo Achille, Konaté Zanga Youssouf, Boere Djibril, Ake Flavien, Sawadogo Guetwendé, Zoma Lamoussa Robert, Meda Nicolas

**Author notes:** **Author corresponding**: Désiré Lucien Dahourou. These authors contributed equally to this work (Kiebré Pegd-Wendé Blaise, Dahourou Desiré Lucien).

## Abstract

**Introduction:** The burden of morbidity and mortality from severe pneumonia remains high among children under five, particularly in resource-limited countries such as Burkina Faso. Targeted hospital-based interventions are essential to achieve Sustainable Development Goal 3.2, which aims to reduce under-five mortality to 25 deaths per 1,000 live births by 2030.

**Methods:** A prospective cohort study was conducted in the pediatrics department of Regional Referral Hospital of Banfora, including 1,406 children aged 2–59 months hospitalized for severe pneumonia between August 2021 and January 2024. Predictors of mortality were identified using the Fine and Gray competing risk model.

**Results:** A total of 1,406 children were included in the analysis. The median age was 18 months (interquartile range: 9–32 months), and 53% were male. During hospitalization, 8.96% (126/1,406; 95% CI: 7.58–10.57) died, corresponding to a mortality rate of 1.93 per 100 person-days. Age < 12 months doubled the instantaneous risk of death. Additionally, hypoxemia (SaO₂ < 90%) (adjusted subdistribution hazard ratio [aSHR]: 1.54; 95% CI: 1.02–2.32), hospitalization during the rainy season (aSHR: 1.73; 95% CI: 1.18–2.54), convulsions (aSHR: 2.93; 95% CI: 1.92–4.47), the presence of stridor (aSHR: 2.17; 95% CI: 1.46–3.22), and hypoglycemia (aSHR: 2.37; 95% CI: 1.45–3.88) increased the risk of death. However, the risk of death was significantly lower in children with moderate or severe anaemia (respectively aSHR = 0.45; 95% CI: 0.24 - 0.84 and aSHR = 0.26; 95% CI: 0.13 - 0.52) and having received antibiotic therapy (ceftriaxone alone [SHRa = 0.52; 95% CI: 0.30 - 0.90] or combined with gentamicin (SHRa = 0.45; 95% CI: 0.30-0.69); ampicillin [aSHR = 0,35; IC95%: 0,13-0,97] and ampicillin combined with gentamicin (SHRa = 0.43; CI: 0.20 - 0.95).

**Conclusion:** The incidence and in-hospital mortality of severe pneumonia are a cause for concern at the Regional Referral Hospital of Banfora and in Burkina Faso. Targeted interventions in hospital settings are necessary and can help achieve Sustainable Development Goal 3.2 on reducing under-five mortality to 25 deaths per 1,000 live births by 2030.

## Introduction

Despite the significant reduction in under-five mortality from 93 deaths per 1000 live births in 1990 to 37 in 2023, child mortality remains high globally, with substantial national and regional disparities [1,2]. While some countries, such as Cabo Verde and South Africa, have already achieved the 2030 Sustainable Development Goal of fewer than 25 deaths of children aged under 59 months per 1,000 live births, projections are not optimistic for low- and middle-income countries (LMICs) such as Burkina Faso. According to the United Nations Interagency Group for Child Mortality Estimation, the infant mortality rate in Burkina Faso was 58.84 per 1,000 live births in 2021, with acute lower respiratory infections (LRIs) ranking as the second leading cause of death. Mortality from LRIs fell from 14.42 per 1,000 live births in 2000 (95% confidence interval [CI95%]: 12.60–16.26) to 10.98 (95% CI = 6.76–17.827) in 2021 [3]. The significant reduction in mortality is the result of a combination of interventions, including the introduction in 2006 of the *Haemophilus influenzae* vaccine and in 2016 of the *Streptococcus pneumoniae* vaccine (PCV 13) in routine childhood vaccination against pneumonia [4,5]. Although pneumonia-related mortality is largely preventable through the implementation of basic strategies including timely referral for management, appropriate use of antibiotics, vaccination and supportive care, identifying the predictors of mortality in children remains a challenge, particularly in LMICs [6].

In view of the significant burden of childhood pneumonia, the World Health Organization (WHO) revised the classification of pneumonia in 2014 into two categories based on simple clinical criteria. One of the aims of this revision was to draw the attention of clinical practitioners and decision-makers to the ‘severe pneumonia’ category because of its high mortality rate [7]. The factors associated with pneumonia-related mortality identified in the literature in children under five years of age in low and middle-income countries (LMICs) are female sex, young age (<12 months), signs of respiratory struggle, altered consciousness, severe anemia, leukocytosis, malnutrition, lack of exclusive breastfeeding, lack of vaccination, and presence of severe illness on admission [8–12].

In Burkina Faso, few data have been reported on inpatient pneumonia-related mortality and its associated factors, despite the fact that it remains the predominant LRI. However, identifying the predictors of mortality is important to guide interventions to reduce the burden of pneumonia-related mortality [8]. This study aimed to estimate mortality rate among children aged 2-59 months hospitalized for severe pneumonia and to identify its risk factors in Burkina Faso.

## Methods

### Study design and framework

We used data from a surveillance study of severe pneumonia among children under five in Burkina Faso, conducted by the Agence de Médecine Préventive – Burkina Faso Office in collaboration with Davycass and funded by the U.S. Centers for Disease Control and Prevention (CDC Atlanta). This was a prospective cohort study, the main objectives of which were, firstly, to assess the impact of the new PCV13 vaccination schedule with a 2-dose primary vaccination plus a booster (2p+1 schedule) on hospitalization of children aged 2-59 months for severe pneumonia. The second aim was to assess the impact of the 2p+1 PCV13 regimen on the prevalence of vaccine-type pneumococcal carriage in children aged 2-59 months hospitalized for severe pneumonia. This was a multicenter study conducted in four public hospitals in Burkina Faso: the Regional Referral Hospital of Banfora in the Cascades region and three first-level referral hospitals located in the health districts of Dô and Dafra (Hauts-Bassins region) and Boulmiougou (Center region). The present analysis used data from the Regional Referral Hospital in Banfora. The Banfora Regional Hospital is a secondary-level referral facility that receives patients referred from health centers across the three districts of the Cascades Region. The other participating sites are first-level referral hospitals that serve patients referred from their respective health areas within their districts. Microbiological samples (nasopharyngeal swabs) were analyzed at the Muraz Center in Bobo Dioulasso.

The Regional Referral Hospital of Banfora is a second-level referral hospital in Burkina Faso, with a catchment area extending to the boundaries of the Cascades region. It covers an area of 18,989 km^2^, bordered to the north by the Hauts-Bassins region, to the east by the South-West region, to the south by Côte d’Ivoire and to the west by Mali. The Cascades health region has a population of 910,642 distributed across three health districts (Banfora, Sindou and Mangodara). The Regional Referral Hospital of Banfora has a capacity of 300 beds, with 196 operational beds, and 12 clinical departments. It is the only medical center in the entire Cascades Health Region capable of offering blood transfusion services. The coverage rate for the third dose of PCV-13 vaccine (Pneumo 3) in the Cascades region was 105.2% in 2022. The pediatric ward where the study was conducted has four units (emergency, hospitalization, neonatology and in-house management of severe acute malnutrition) with a capacity of 50 beds. During the data collection period, the department had two pediatricians, 10 general practitioners and 52 nurses. The doctors were responsible for admitting patients and monitoring them daily. Outpatient consultations were conducted to monitor patients after hospitalization.

From August 30, 2021, to January 31, 2024, hospitalized children aged 2–59 months who met the WHO criteria for severe pneumonia were recruited into the study and followed up during their hospital stay. Severe pneumonia is defined by the WHO as the presence of cough and/or difficulty breathing that has been evolving for less than two weeks with one or more general danger signs. Difficulty breathing was defined as chest retraction or rapid breathing (respiratory rate ≥ 50 breaths per minute in a child aged 2 to < 12 months or ≥ 40 breaths per minute in a child ≥ 12 months) in a child without wheezing. Children with wheezing should be reassessed two hours after bronchodilator administration. General danger signs include inability to drink or breastfeed, persistent vomiting, convulsions, lethargy or loss of consciousness, stridor in a calm child, and severe malnutrition [7].

### Data collection

Data were collected using paper-based case report forms (CRFs) and then entered into the STELab electronic platform (système de transport des échantillons de laboratoire) dedicated to monitoring and tracking the transport of biological samples of infectious diseases under surveillance in Burkina Faso.

Sociodemographic characteristics and disease history were collected in the CRF using a structured questionnaire administered directly by the investigators to the parents or legal guardians of the children. Clinical and diagnostic information, vaccination status and outcomes were collected by direct administration of a structured questionnaire to the parents or legal guardians of the children, including sociodemographic variables and medical history. Clinical, diagnostic, outcome and vaccination status information were also obtained from the children’s medical records and health diaries. Only clinical and diagnostic characteristics on admission to the pediatric emergency department were considered.

### Study variables

The dependent variable was in-hospital mortality rate. Independent variables included sociodemographic, anthropometric, environmental, clinical, paraclinical, and therapeutic parameters. Sociodemographic characteristics included the child’s age, categorized into four groups (2–5 months, 6–11 months, 12–23 months, and 24–59 months), and sex (male or female). Anthropometric indicators comprised mid-upper arm circumference (MUAC), categorized as <115 mm, 115–124 mm, or ≥125 mm, as well as z-scores for weight-for-height (WHZ), weight-for-age (WAZ), and height-for-age (HAZ), classified as < -3, between -3 and -2, or ≥ -2. Severe acute malnutrition was defined as any of the following: WHZ < -3, MUAC <115 mm, or clinical signs suggestive of malnutrition, such as bilateral edema. For infants under six months, additional criteria included being too weak to breastfeed, failure to gain weight, or losing 10% or more of their initial body weight [13]. Environmental factors included the season of hospitalization, defined as the dry season (November to May) and rainy season (June to October), as well as reported household exposure to secondhand smoke and the use of fumigating substances (e.g., incense and mosquito coils) in the child’s bedroom.

Clinical variables collected at admission included signs of pneumonia, such as cough and an age-specific elevated respiratory rate, as well as danger signs, including inability to drink or breastfeed, persistent vomiting, convulsions, lethargy, altered consciousness, stridor at rest, subcostal chest retractions or signs of severe acute malnutrition. Axillary temperature was categorized as hypothermia (<36°C), normal (36 to <38°C), or fever (≥38°C). Heart rate was classified into three categories (normal, bradycardia, and tachycardia), whereas respiratory rate was categorized as either normal or tachypnea according to age-specific thresholds [14]. Hypoxemia was defined as a peripheral oxygen saturation < 90% in room air. Vaccination status against Streptococcus pneumoniae (PCV13) and Haemophilus influenzae b (Hib) was assessed and categorized as full vaccine, incomplete vaccine, or no vaccine.

Biological diagnostic variables included the white blood cell (WBC) count, categorized as leukopenia (<4,000/mm³), normal (4,000–10,000/mm³), or leukocytosis (>10,000/mm³); hemoglobin concentration, classified as severe anemia (<7 g/dL), moderate anemia (7 to <12 g/dL), or normal (≥12 g/dL); and blood glucose level, categorized as hypoglycemia (<2.7 mmol/L) or normal (≥2.7 mmol/L). The presence of malaria was assessed using a rapid diagnostic test or thick blood smear. Initial antibiotic therapy was classified based on the administered molecule or combination: ceftriaxone, gentamicin, ampicillin, ceftriaxone + gentamicin, or gentamicin + ampicillin. Length of hospital stay was categorized as 1, 2–3, 4–7, or > 7 days. Discharge outcomes were grouped into three categories: recovered, deceased, or referred/discharged against medical advice.

### Statistical Analysis

Categorical variables were described using frequencies and percentages, and comparisons between groups were performed using the chi-square test or Fisher’s exact test, as appropriate. Continuous variables were summarized using medians and interquartile ranges (IQR), and group comparisons were performed using the Wilcoxon rank-sum test. The incidence of death during hospitalization was estimated at 100 person-days. Follow-up time was calculated from the date of admission to the date of discharge alive, referral, death or discharge against medical advice.

To identify predictors of mortality, we used a competing risk survival model based on the Fine and Gray method subdistribution hazard approach [15]. In this analysis, the event of interest was in-hospital death, which was analyzed in competition with the events “referred” and “discharged against medical advice (DAMA)” The category “referred” included all patients who were transferred to the Souro Sanou University Hospital Center for further management. Patients who were discharged alive, referred or discharged against medical advice were censored at the date of discharge or last contact.In univariable analysis, variables associated with death at the 20% threshold were retained for multivariable analysis. A backward stepwise selection strategy was applied to identify independent predictors of mortality at the 5% significance level. All analyses were performed using Stata version 16 (StataCorp).

### Ethical Considerations

The severe pneumonia hospitalization surveillance study received ethical approval from the National Health Research Ethics Committee (approval number 2020-02-021). Authorization for the use of these data was granted by the Ministry of Health through official note N°2024-3660/MSHP/SG/DGESS/DSSE dated June 16, 2024, and by the Director General of the Regional Referral Hospital of Banfora through note No. 2024-31/MSHP/SG/CHR-BFR/DG dated June 26, 2024. Data were extracted anonymously from the Laboratory Sample Transport System (STELab) database.

## Results

Between August 30, 2021, and January 31, 2024, 1,579 children aged 2–59 months were admitted with severe pneumonia, accounting for 25.36% (1,579/6,226) of all hospitalizations in this age group. Of these, 1,406 (89.04%) were enrolled in the study. Overall, 8.96% (126/1,406) of patients died during hospitalization, and 4.84% (68/1,406) were either referred or discharged against medical advice (Figure 1).

### Sociodemographic and anthropometric characteristics

The median age of participants was 18 months (IQR: 9–32 months). More than half of the participants were male (53.20%, 748/1406), and discharge outcomes did not significantly differ by sex (p = 0.205). In addition, 19.77% (278/1406) had severe stunting and 24.32% (342/1406) were severely underweight (WAZ < -3). Discharge outcomes varied significantly by age group (p = 0.000) and season of hospitalization (p = 0.014) (Table 1).

**Table 1.** Sociodemographic and anthropometric characteristics of children aged 2 to 59 months hospitalized with severe pneumonia at the Regional Referral Hospital of Banfora, from August 30, 2021, to January 31, 2024, according to discharge outcome

| Variables | Outcomes | | | p-value ($\chi^2$ ou Fisher) | Total (%) N = 1406 |
| --- | --- | --- | --- | --- | --- |
|  | Recovered n(%) | Deceased n(%) | Referred or DAMA n (%) |  |  |
| **Sex** |  |  |  | **0.205** |  |
| Female | 565 (46.58) | 55 (43.65) | 38 (56.72) |  | 658 (46.80) |
| Male | 648 (53.42) | 71 (56.35) | 29 (43.28) |  | 748 (53.20) |
| **Median age in months (IQR)** | **19 (10-33)** | **11.5 (5-25)** | **13 (6-24)** |  | **18 (9-32)** |
| **Age (months)** |  |  |  | **0.000** |  |
| [2 –5[ | 172 (14.18) | 33 (26.19) | 12 (17.91) |  | 217 (15.43) |
| [6 – 11[ | 206 (16.98) | 30 (23.81) | 16 (23.88) |  | 252 (17.92) |
| [12 – 23[ | 308 (25.39) | 27 (21.43) | 19 (28.36) |  | 354 (25.18) |
| [24 – 59 [ | 527 (43.45) | 36 (28.57) | 20 (29.85) |  | 583 (41.47) |
| **Hospitalization season** |  |  |  | **0.014** |  |
| Dry | 549 (45.26) | 40 (31.75) | 31 (46.27) |  | 620 (44.10) |
| Rainy | 664 (54.74) | 86 (68.25) | 36 (53.73) |  | 786 (55.90) |
| **Median WHZ (IQR)** | **-1.72 (-3.16 à -0.28)** | **-1.94 (-3.38 à -0.38)** | **-2.07 (-3.52 à -0.05)** |  | **-1.77 (-3.20 to -0.28)** |
| **WHZ** |  |  |  | **0.406** |  |
| WHZ ≥ -2 | 674 (55.56) | 64 (50.79) | 31 (46.27) |  | 769 (54.69) |
| -3 ≤ WHZ < -2 | 204 (16.82) | 22 (17.46) | 16 (23.88) |  | 242 (17.21) |
| WHZ < -3 | 335 (27.62) | 40 (31.75) | 20 (29.85) |  | 395 (28.09) |
| **Median HAZ (IQR)** | **-1.15 (-2.49 à 0.17)** | **-1.20 (-2.29 à 0.35)** | **-1.40 (-2.72 à 0.29)** |  | **-1.17 (-2.47 to 0.17)** |
| **HAZ** |  |  |  | **0.555** |  |
| HAZ ≥ -2 | 808 (66.61) | 93 (73.81) | 44 (65.67) |  | 945 (67,21) |
| -3 ≤ HAZ < -2 | 159 (13.11) | 14 (11.11) | 10 (14.93) |  | 183 (13,02) |
| HAZ < -3 | 246 (20.28) | 19 (15.08) | 13 (19.40) |  | 278 (19,77) |
| **Median WAZ (IQR)** | **-1.72 (-2.93 à -0.69)** | **-1.79 (-3.05 à -0.90)** | **-2.03 (-2.91 à -1.15)** |  | **-1.75 (-2.93 to -0.74)** |
| **WAZ** |  |  |  | **0.456** |  |
| WAZ ≥ -2 | 687 (56.64) | 69 (54.76) | 33 (49.25) |  | 789 (56.12) |
| -3 ≤ WAZ < -2 | 231 (19.04) | 25 (19.84) | 19 (28.36) |  | 275 (19.56) |
| WAZ < -3 | 295 (24.32) | 32 (25.40) | 15 (22.39) |  | 342 (24.32) |
| **Median MUAC (IQR)** | **13.5 (12.5 à 14)** | **13 (11.6 à 13.8)** | **13 (12.2 à 13.8)** |  | **13.5 (12.4 to 14)** |
| **MUAC** |  |  |  | **0.044** |  |
| ≥ 125 mm | 917 (75.60) | 87 (69.05) | 49 (73.13) |  | 1053 (74.89) |
| 115 ≤ MUAC < 125 mm | 119 (9.81) | 9 (7.14) | 4 (5.97) |  | 132 (9.39) |
| MUAC < 115 mm | 177 (14.59) | 30 (23.81) | 14 (20.90) |  | 221 (15.72) |

IQR: medians with interquartile ranges; WHZ: weight-for-height z-score; MUAC: mid-upper arm circumference; HAZ: height-for-age z-score; WAZ: weight-for-age z-score; DAMA: discharged against medical advice

### Clinical characteristics

At admission, 45.38% (638/1,406) of the children were febrile and 72.97% (1,026/1,406) presented with tachypnea. The most common danger signs were altered consciousness (80.23%, 1,128/1,406), subcostal chest retraction (39.26%, 552/1,406), and stridor (20.20%, 285/1,406), all of which varied significantly by discharge outcome. Regarding vaccination status, 65.63% (693/1,056) were up to date with their immunization against *Streptococcus pneumoniae* (PCV13) and *Haemophilus influenzae*. The median length of hospital stay was 4 days (IQR: 3–6 days) (Table 1).

**Table 2.** Clinical Characteristics of Children Aged 2 to 59 Months Hospitalized with Severe Pneumonia at the Regional Referral Hospital of Banfora, from August 30, 2021, to January 31, 2024

| Variables | Outcomes | | | p-value ($\chi^2$ ou Fisher) | Total (%) N = 1406 |
| --- | --- | --- | --- | --- | --- |
|  | Recovered n(%) | Deceased n(%) | Referred or DAMA n (%) |  |  |
| **Temperature** |  |  |  | **0.103** |  |
| Apyrexie | 634 (52.27) | 51 (40.48) | 30 (44.78) |  | 715 (50.85) |
| Hypothermie | 45 (3.71) | 6 (4.76) | 2 (2.99) |  | 53 (3.77) |
| Hyperthermie | 534 (44.02) | 69 (54.76) | 35 (52.24) |  | 638 (45.38) |
| **SaO2** |  |  |  | **0.000** |  |
| Normal | 1047 (86.31) | 77 (61.11) | 53 (79.10) |  | 1177 (83.71) |
| Low | 166 (13.69) | 49 (38.89) | 14 (20.90) |  | 229 (16.29) |
| **Median respiration rate (IQR)** | **46 (42-52)** | **53.5 (45-64)** | **47 (43-52)** |  | **47 (42-54)** |
| **Respiratory rate** |  |  |  | **0.019** |  |
| Normal | 338 (27.86) | 21 (16.67) | 21 (31.34) |  | 380 (27.03) |
| Tachypnea | 875 (72.14) | 105 (83.33) | 46 (68.66) |  | 1026 (72.97) |
| **Median Heart rate (IQR)** | **148 (133-165)** | **151 (132-173)** | **161 (142-172)** |  | **149.5 (134-167)** |
| **Heart rate** |  |  |  | **0.005** |  |
| Normal | 565 (46.58) | 53 (42.06) | 21 (31.34) |  | 639 (45.45) |
| Bradycardia | 42 (3.46) | 11 (8.73) | 2 (2.99) |  | 55 (3.91) |
| Tachycardia | 606 (49.96) | 62 (49.21) | 44 (65.67) |  | 712 (50.64) |
| **Tabacco exposure** |  |  |  | **0.560** |  |
| Yes | 77 (6.35) | 11 (8.73) | 4 (5.97) |  | 92 (6.54) |
| No | 1136 (93.65) | 115 (91.27) | 63 (94.03) |  | 1314 (93.46) |
| **Use fumigene in room** |  |  |  | **0.471** |  |
| No | 157 (12.94) | 12 (9.52) | 10 (14.93) |  | 179 (12.73) |
| Yes | 1056 (87.06) | 114 (90.48) | 57 (85.07) |  | 1227 (87.27) |
| **Number of danger signs** |  |  |  | **0.000** |  |
| One sign | 493 (40.64) | 27 (21.43) | 25 (37.31) |  | 545 (38.76) |
| 2 signs | 527 (43.45) | 53 (42.06) | 27 (40.30) |  | 607 (43.17) |
| 3 or more | 193 (15.91) | 46 (36.51) | 15 (22.39) |  | 254 (18.07) |
| **Inability to drink or breastfeed** |  |  |  | **0.002** |  |
| No | 1115 (91.92) | 105 (83.33) | 57 (85.07) |  | 1277 (90.83) |
| Yes | 98 (8.08) | 21 (16.67) | 10 (14.93) |  | 129 (9.17) |
| **Severe acute malnutrition** |  |  |  | **0.963** |  |
| No | 860 (70.90) | 88 (69.84) | 47 (70.15) |  | 995 (70.77) |
| Yes | 353 (29.10) | 38 (30.16) | 20 (29.85) |  | 411 (29.23) |
| **Stridor** |  |  |  | **0.000** |  |
| No | 988 (81.45) | 77 (61.11) | 57 (85.07) |  | 1122 (79.80) |
| Yes | 225 (18.55) | 49 (38.89) | 10 (14.93) |  | 284 (20.20) |
| **Persistent vomiting** |  |  |  | **0.182** |  |
| No | 839 (69.17) | 97 (76.98) | 48 (71.64) |  | 984 (69.99) |
| Yes | 374 (30.83) | 29 (23.02) | 19 (28.36) |  | 422 (30.01) |
| **Convulsion** |  |  |  | **0.000** |  |
| No | 937 (77.25) | 74 (58.73) | 47 (70.15) |  | 1058 (75.25) |
| Yes | 276 (22.75) | 52 (41.27) | 20 (29.85) |  | 348 (24.75) |
| **Lethargy or unconsciousness** |  |  |  | **0.377** |  |
| No | 247 (20.36) | 20 (15.87) | 11 (16.42) |  | 278 (19.77) |
| Yes | 966 (79.64) | 106 (84.13) | 56 (83.58) |  | 1128 (80.23) |
| **Cough** |  |  |  | **0.046** |  |
| No | 247 (20.36) | 36 (28.57) | 10 (14.93) |  | 293 (20.84) |
| Yes | 966 (79.64) | 90 (71.43) | 57 (85.07) |  | 1113 (79.16) |
| **Chest retractions** |  |  |  | **0.000** |  |
| No | 763 (62.90) | 48 (38.10) | 43 (64.18) |  | 854 (60.74) |
| Yes | 450 (37.10) | 78 (61.90) | 24 (35.82) |  | 552 (39.26) |
| **Vaccinal statut** |  |  |  | **0.148** |  |
| Full vaccine | 596 (65.49) | 61 (62.89) | 36 (73.47) |  | 693 (65.63) |
| Infull vaccine | 260 (28.57) | 24 (24.74) | 11 (22.45) |  | 295 (27.94) |
| No vaccine | 54 (5.93) | 12 (12.37) | 2 (4.08) |  | 68 (6.44) |
| **Median length of stay (IQR)** | **4 (3-6)** | **2 (1-4)** | **2 (1-5)** |  | **4 (3-6)** |
| **Length of stay** |  |  |  | **0.000** |  |
| 1 day | 40 (3.30) | 38 (30.16) | 26 (38.81) |  | 104 (7.40) |
| 2 – 3 days | 513 (42.29) | 55 (43.65) | 14 (20.90) |  | 582 (41.39) |
| 4 - 7 days | 475 (39.16) | 23 (18.25) | 21 (31.34) |  | 519 (36.91) |
| > 7 days | 185 (15.25) | 10 (7.94) | 6 (8.96) |  | 201 (14.30) |

IQR: medians with interquartile ranges; DAMA: discharged against medical advice; SaO<sub>2</sub>: oxygen peripheral saturation

### Diagnostic and therapeutic characteristics

Malaria co-infection was present in 59.20% (827/1,406) of participants. Nearly all children presented with either moderate or severe anemia, showing significant variation by discharge outcome (p = 0.000). Hypoglycemia occurred in 8.17% (113/1,383) of participants, also reflecting a statistically significant difference based on discharge status (p = 0.000). From a therapeutic perspective, antibiotics use during hospitalization varied significantly by discharge outcome (p = 0.002). The most frequently administered antibiotics were ceftriaxone (21.28%; 300/1,406) and the combination of ceftriaxone and gentamicin (31.72%; 446/1,406). (Table 3)

**Table 3.** Diagnostic and therapeutic characteristics of children aged 2 to 59 months hospitalized with severe pneumonia at the Regional Referral Hospital of Banfora, from August 30, 2021, to January 31, 2024

| Variables | Outcomes | | | p-value<br>($\chi^2$ ou<br>Fisher) | Total (%) N =<br>1406 |
| --- | --- | --- | --- | --- | --- |
|  | Recovered<br>n (%) | Deceased<br>n (%) | Referred<br>or DAMA n<br>(%) |  |  |
| **Malaria** |  |  |  | **0.076** |  |
| Non | 487 (40.41) | 61 (48.80) | 22 (32.84) |  | 570 (40.80) |
| Oui | 718 (59.59) | 64 (51.20) | 45 (67.16) |  | 827 (59.20) |
| **Median white blood cell**<br>**count (IQR)** | 12745<br>(8910–<br>19060) | 13945<br>(8315–<br>22440) | 14010<br>(9410–<br>21620) |  | 12980 (8900–<br>19440) |
| **White blood cell count** |  |  |  | **0.241** |  |
| Normal | 354 (29.18) | 30 (23.81) | 13 (19.40) |  | 397 (28.24) |
| Leukopenia | 45 (3.71) | 7 (5.56) | 4 (5.97) |  | 56 (3.98) |
| Leukocytosis | 814 (67.11) | 89 (70.63) | 50 (74.63) |  | 953 (67.78) |
| **Median hemoglobin**<br>**concentration (IQR) (g/dL)** | 7.2 (4.8-9.3) | 8.1 (6.15-9.8) | 4.8 (3.1-8.3) |  | 7.3 (4.8-9.35) |
| **Hemoglobin concentration**<br>**(g/dL)** |  |  |  | **0.000** |  |
| Normal ($\geq 12$) | 38 (3.13) | 11 (8.73) | 1 (1.49) | | 50 (3.56) |
| Moderate anemia (7–12) | 601 (49.55) | 75 (59.52) | 20 (29.85) |  | 696 (49.50) |
| Severe anemia ($\leq 7$) | 574 (47.32) | 40 (31.75) | 46 (68.66) | | 660 (46.94) |
| **Median glycemia (IQR)** | 5.68 (4.63-6.99) | 5.69 (3.25-8.05) | 5.18 (4.56-6.2) |  | 5.67 (4.57-7) |
| **Glycemia** |  |  |  | **0.000** |  |
| Normal | 1114 (93.22) | 97 (78.86) | 59 (90.77) |  | 1270 (91.83) |
| Hypoglycemia | 81 (6.78) | 26 (21.14) | 6 (9.23) |  | 113 (8.17) |
| **Type of antibiotic** |  |  |  | **0.002** |  |
| Aucun | 364 (30.01) | 58 (46.03) | 28 (41.79) |  | 450 (32.01) |
| Ampicillin | 82 (6.76) | 4 (3.17) | 4 (5.97) |  | 90 (6.40) |
| Ampicillin.Gentamicin | 108 (8.90) | 10 (7.94) | 4 (5.97) |  | 122 (8.68) |
| Ceftriaxone | 267 (22.01) | 17 (13.49) | 11 (16.42) |  | 295 (20.98) |
| Ceftriaxone.Gentamicin | 391 (32.23) | 36 (28.57) | 19 (28.36) |  | 446 (31.72) |
| Gentamicin | 1 (0.08) | 1 (0.79) | 1 (1.49) |  | 3 (0.21) |

*IQR: medians with interquartile ranges; DAMA: discharged against medical advice*

### Overall mortality

The median follow-up time for the 1,406 participants was 6,540 person-days. During this period, 126 children (8.96%; IC95%: 7,58-10,57) died. The overall mortality rate was 1.93 per 100 person-days (95% CI: 1.62 to 2.29). The probability of death was 2.7% on the first day of hospitalization, cumulative probability of death is 7.01% at the third day, and 11.02% after seven days of hospitalization (Figure 2).

### Predictors of mortality in severe pneumonia

In univariable analysis (Table 4), 21 variables were associated with mortality at a 20% significance threshold.

**Table 4.**
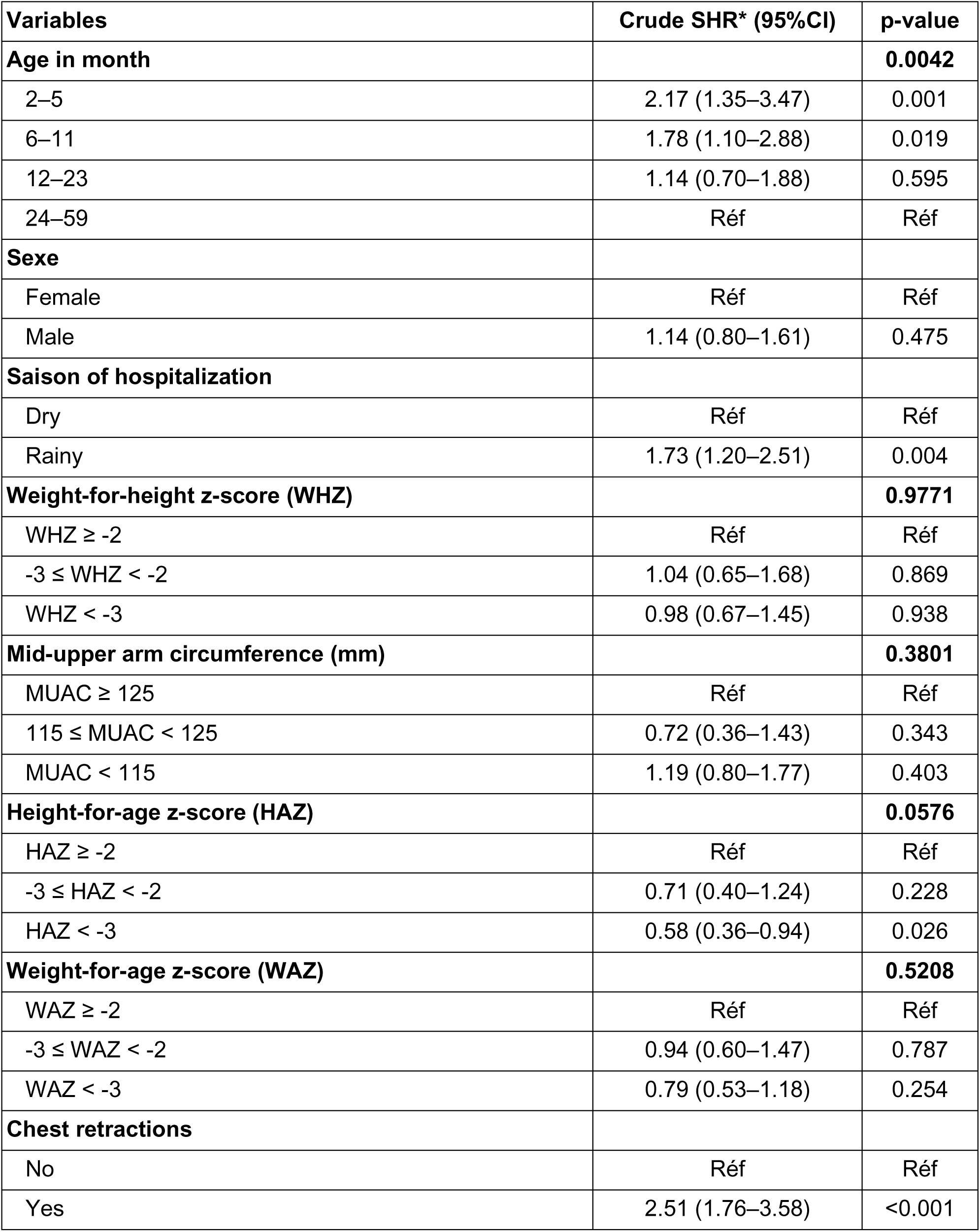
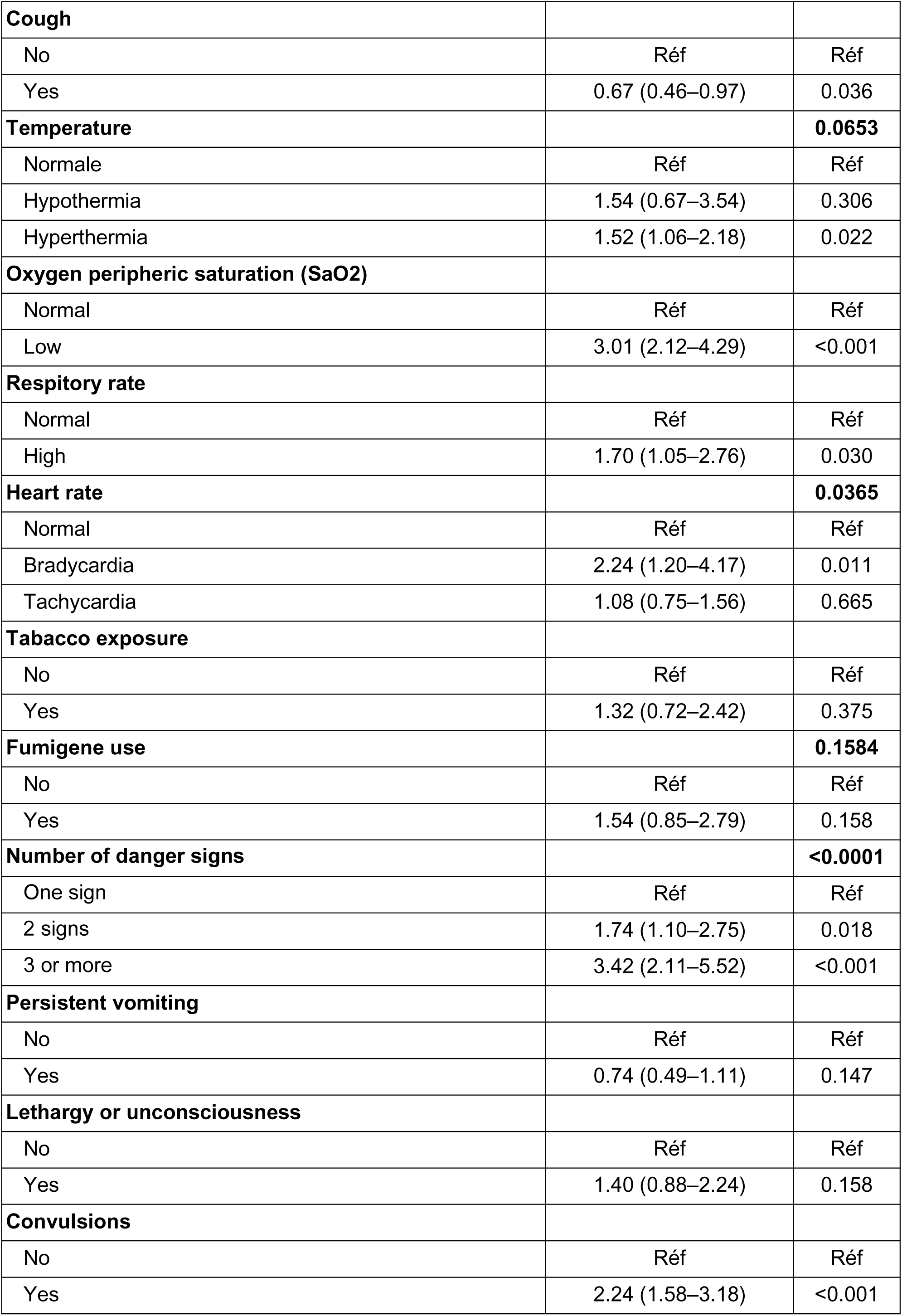
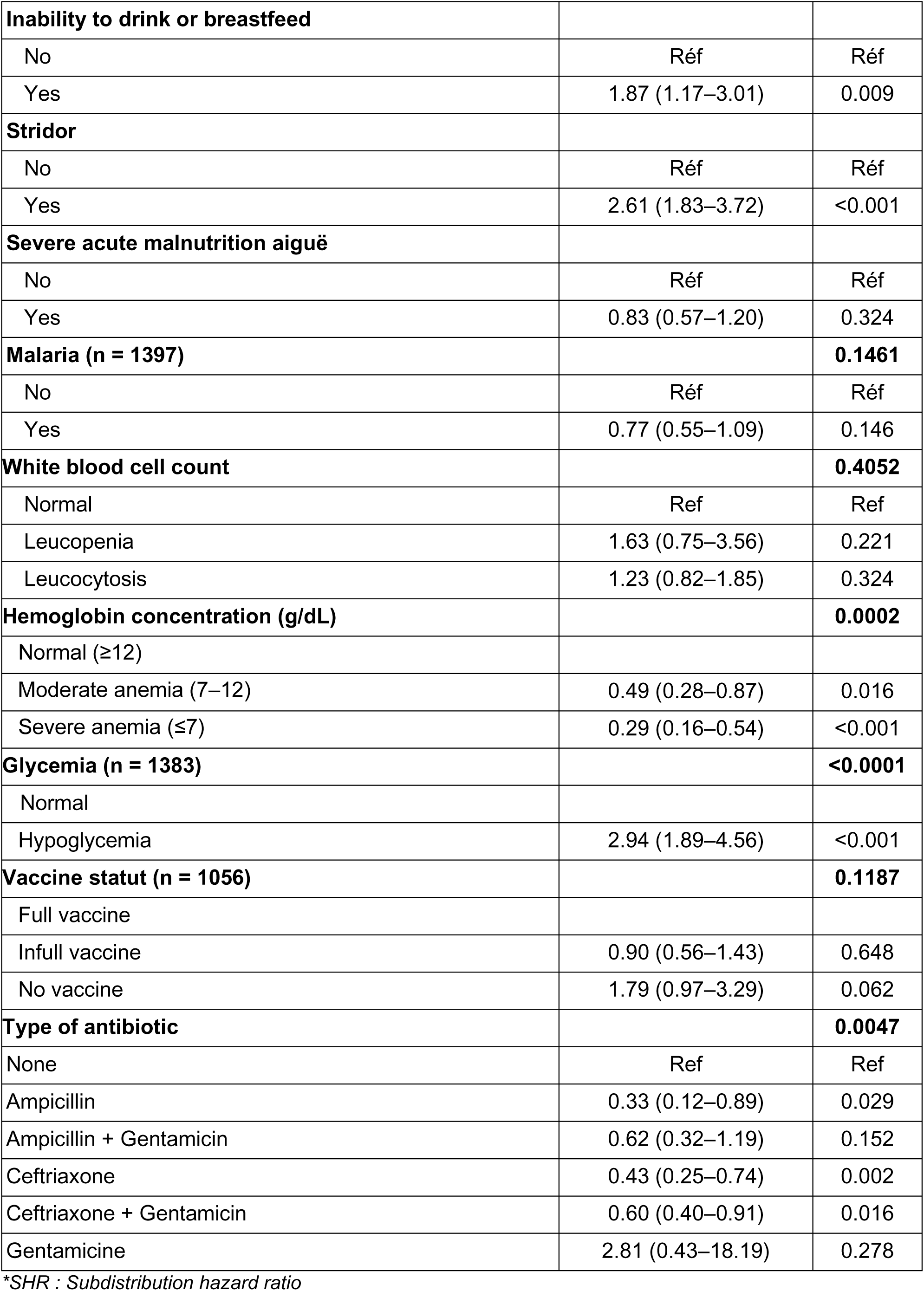
Factors associated with mortality among children aged 2 to 59 months hospitalized with severe pneumonia at the Regional Referral Hospital of Banfora, from August 30, 2021, to January 31, 2024 – univariable competing risk regression analysis

In the multivariable competing risks final model, nine variables included age, season of hospitalization, hypoxemia, bradycardia, stridor, convulsions, hypoglycemia, type of antibiotic therapy and hemoglobin concentration, were independently associated with the instantaneous risk of death. Compared to children aged 24–59 months, those aged 2-5 months and 6-11 months had a significantly higher risk of mortality, with adjusted subdistribution hazard ratios (SHRs) of 2,28 (1,28-4,07) and 2.02 (95% CI: 1.23–3.36), respectively. Similarly, the presence of convulsions and stridor was associated with increased mortality risk, with adjusted SHRs of 2.93 (95% CI: 1.92-4.47) and 2.17 (95% CI: 1.46-3.22), respectively. (Table 5).

**Table 5.** Factors Associated with Mortality Among Children Aged 2 to 59 Months Hospitalized with Severe Pneumonia at the Regional Referral Hospital of Banfora (August 30, 2021 – January 31, 2024): Multivariable Fine and Gray Competing Risk Regression

| Variable | Full model |  | Final model |  |
| --- | --- | --- | --- | --- |
|  | Ajusted SHR* (IC à 95 %) | p-value | Ajusted SHR (IC à 95 %) | p-value |
| **Age (n=1380)** |  | **0.0461** |  | **0.020** |
| 24-59 | Ref |  | Ref |  |
| 2-5 mois | 2.36 (1.25–4.45) | 0.008 | 2.28 [1.28-4.07] | 0.005 |
| 6-11 mois | 2.08 (1.13–3.81) | 0.018 | 2.02 [1.22-3.36] | 0.007 |
| 12-23 mois | 1.41 (0.83–2.40) | 0.202 | 1.47 [0.88-2.48] | 0.144 |
| **Saison of hospitalization (n=1380)** |  |  |  |  |
| Dry | Ref |  | Ref |  |
| Rainy | 1.73 (1.19–2.51) | 0.004 | 1.73 [1.18-2.54] | 0.005 |
| **Height-for-age z-score (HAZ) (n=1380)** |  | **0.1445** | --- | --- |
| HAZ≥-2 | Ref | | --- | --- |
| -3 ≤ HAZ < -2 | 0.93 (0.53–1.65) | 0.814 | --- | --- |
| HAZ < -3 | 0.58 (0.34–1.00) | 0.049 | --- | --- |
| **Chest retraction** |  |  | --- | --- |
| No | Ref |  | --- | --- |
| Yes | 1.22 (0.77–1.93) | 0.390 | --- | --- |
| **Cough** |  |  | --- | --- |
| No | Ref |  | --- | --- |
| Yes | 0.68 (0.44–1.04) | 0.078 | --- | --- |
| **Temperature** |  | **0.0794** | --- | --- |
| Normal | Ref |  | --- | --- |
| Hypothermia | 2.10 (0.99–4.48) | 0.054 | --- | --- |
| Hyperthermia | 1.37 (0.92–2.05) | 0.119 | --- | --- |
| **Oxygen peripheric saturation** |  |  | --- | --- |
| Normal | Ref |  | --- | --- |
| Low (SaO<sub>2</sub><90%) | 1.29 (0.84–1.99) | 0.239 | 1.54 [1.02-2.32] | 0.039 |
| **Respirator rate** |  |  |  |  |
| Normal | Ref |  | Ref |  |
| High | 1.69 (0.99–2.90) | 0.056 | 1.80 (1.06–3.05) | 0.029 |
| **Heart rate** |  | **0.0479** |  | **0.114** |
| Normal | Ref |  | Ref |  |
| Bradycardia | 2.11 (1.12–3.95) | 0.020 | 1.96 [1.04-3.69] | 0.037 |
| Tachycardia | 1.03 (0.68–1.56) | 0.897 | 1.14 [0.77-1.69] | 0.513 |
| **Fumigene use** |  |  | --- | --- |
| No | Ref |  | --- | --- |
| Yes | 1.72 (0.87–3.42) | 0.119 | --- | --- |
| **Number of danger signs** |  | **0.8284** | --- | --- |
| 1 sign | Ref |  | --- | --- |
| 2 signs | 1.19 (0.60–2.35) | 0.616 | --- | --- |
| ≥3 signs | 1.40 (0.48–4.10) | 0.540 | --- | --- |
| **Persistent vomiting** |  |  | --- | --- |
| No | Ref |  | --- | --- |
| Yes | 0.98 (0.51–1.90) | 0.959 | --- | --- |
| **Lethargy or unconsciousness** |  |  | --- | --- |
| No |  |  | --- | --- |
| Yes | 0.92 (0.46–1.84) | 0.808 | --- | --- |
| **Convulsion** |  |  |  |  |
| No | Ref |  | Ref |  |
| Yes | 2.42 (1.34–4.34) | 0.003 | 2.93 (1.92–4.47) | <0.001 |
| **Inability to drink or breastfeed** |  |  | --- | --- |
| No | Ref |  | --- | --- |
| Yes | 1.56 (0.83–2.92) | 0.167 | --- | --- |
| **Stridor** |  |  |  |  |
| No | Ref |  | Ref |  |
| Yes | 1.52 (0.84–2.76) | 0.165 | 2.17 (1.46–3.22) | <0.001 |
| **Malaria** |  |  |  |  |
| No | Ref |  | --- | --- |
| Yes | 0.78 (0.47–1.30) | 0.342 | --- | --- |
| **Hemoglobin concentration** |  | **0.0015** |  | **<0.001** |
| Normal | Ref |  | Ref |  |
| Moderate anemia | 0.41 (0.21–0.80) | 0.009 | 0.45 [0.24-0.84] | 0.012 |
| Severe anemia | 0.27 (0.13–0.55) | <0.001 | 0.26 [0.13-0.52] | <0.001 |
| **Glycemia** |  |  |  |  |
| Normal | Ref |  | Ref |  |
| Hypoglycemia | 2.50 (1.51–4.14) | <0.001 | 2.37 (1.45–3.88) | <0.001 |
| **Type of antibiotic** |  | **0.0002** |  | **<0.001** |
| None | Ref |  | Ref |  |
| Ampicillin | 0.41 (0.15–1.09) | 0.072 | 0.35 [0.13-0.97] | 0.044 |
| Ampicillin + Gentamicin | 0.44 (0.20–0.97) | 0.043 | 0.43 [0.20-0.95] | 0.036 |
| Ceftriaxone | 0.49 (0.28–0.85) | 0.012 | 0.52 [0.30-0.90] | 0.019 |
| Ceftriaxone + Gentamicin | 0.44 (0.29–0.68) | <0.001 | 0.45 [0.30-0.69] | <0.001 |
| Gentamicin | 2.74 (1.04–7.19) | 0.040 | 1.80 [0.85-3.82] | 0.123 |

\*SHR : Subdistribution hazard ratio

## Discussion

Our study aimed to estimate in-hospital mortality rate associated with severe pneumonia among children aged 2 to 59 months at the regional referral hospital of Banfora in Burkina Faso and identify its predictors. We found that the in-hospital mortality rate remained relatively high (1.93 per 100 person-days) Mortality was significantly higher among children under five years of age who were admitted during the rainy season and who presented with convulsions, stridor, hypoxemia, or hypoglycemia at admission. In contrast, the risk of death was significantly lower among children who received antibiotic treatment and who presented with moderate or severe anemia. However, the reported mortality rate may be an underestimate. Indeed, among the eligible children who were not enrolled in the cohort, 34.32% died. This high pre-inclusion mortality suggests that the cohort primarily included less severely ill children.

This relatively high in-hospital mortality may be explained by the referral status of the Regional Referral Hospital of Banfora. As the only secondary-level referral facility in the Cascades region, it primarily admits patients in serious or critical condition referred from peripheral health centers. This referral pathway can lead to delays in timely and appropriate care, potentially worsening the clinical status of patients, especially since first-level facilities often lack the technical capacity and skilled personnel required for managing severe illnesses. The critical role of delayed management due to inadequate primary care at the initial point of contact in the mortality of severe pneumonia has already been documented in India [16].

Children under one year of age had a significantly higher risk of death. This increased vulnerability is primarily attributed to the immaturity of the immune system at this age and the anatomical and functional immaturity of the respiratory tract, which compromises the ability to mount an effective response to infections and leads to a higher risk of poor outcomes [16]. The association between young age and mortality has been consistently reported in previous studies [17,18]. These findings underscore the importance of early recognition and prompt treatment of severe pneumonia in infants to reduce mortality.

Hospitalization during the rainy season significantly increased the risk of death by nearly 73%. Humid and hot conditions during this period create a favorable environment for the proliferation of microorganisms responsible for lower respiratory tract infections [19]. In addition, the rainy season coincides with a peak in malaria transmission and pediatric hospital admissions. This seasonal overcrowding can compromise the quality of care by delaying timely treatment or limiting close monitoring of critically ill patients, thereby worsening their condition and contributing to excess mortality. Concurrently, the resurgence of malaria during this period may complicate the diagnosis of pneumonia, as the clinical features of severe malaria often overlap with those of severe pneumonia. These two conditions can also coexist, potentially acting synergistically to increase mortality among affected patients [20–22]. However, in our study, malaria co-infection was not identified as a significant risk factor for death, as reported in other studies [20,23].

Hypoxemia is a critical clinical indicator in severe illness. An insufficient supply of oxygen to tissues, if not corrected, can lead to multiorgan failure and death. Meta-analyses have reported that hypoxemia (SpO₂ ≤ 90%) increases the risk of death by fivefold in children with pneumonia [24]. In our study, the risk of death was 54% higher among participants with peripheral oxygen saturation below 90% on room air.

In our study, stridor increased the instantaneous risk of death by more than twofold. This abnormal inspiratory sound typically indicates upper airway obstruction, which may result from severe inflammation or excessive mucus production. The elevated mortality risk observed in children with stridor could therefore reflect respiratory failure due to reduced airway diameter. This association has been reported in previous studies conducted in Malawi [17,25]. These children should be identified early and managed with regular nasopharyngeal suctioning to reduce the risk of hypoxia caused by airway obstruction.

Children presenting with convulsions had nearly a threefold increase in the instantaneous risk of death compared to those without. Convulsions are recognized as clinical danger signs requiring prompt and specialized attention [7]. Several studies in Asia have also identified convulsions as a predictor of death in severe pneumonia [6,26]. A potential mechanism is respiratory distress due to dysfunction of autonomic centers in the medulla oblongata, which may be exacerbated by the use of benzodiazepines for curative or prophylactic sedation—thereby further increasing the risk of mortality [12]. In resource-limited settings such as Burkina Faso, phenobarbital is commonly used for recurrent convulsions. However, caregivers should be made aware of the need for cautious administration in cases of severe pneumonia to avoid aggravating respiratory compromise.

Unexpectedly, moderate or severe anemia was identified as a protective factor in our study. This finding contrasts with previous research that has consistently identified anemia as a major contributor to mortality across various diseases, particularly in cases of severe pneumonia. Anemia reduces the oxygen-carrying capacity of hemoglobin, thereby impairing oxygen delivery to tissues and increasing the risk of respiratory failure which can lead to death [27]. One possible explanation for our results is that anemia was classified based on the initial blood count performed at admission. Children diagnosed with moderate or severe anemia typically received prompt transfusion with packed red blood cells or were referred to another facility in cases where blood products were unavailable. The apparent protective effect of anemia may therefore reflect the beneficial impact of early blood transfusion. Furthermore, transfused children may have received closer clinical attention due to post-transfusion hemovigilance protocols which require enhanced documentation and monitoring. This additional surveillance may have enabled the earlier identification and management of clinical deterioration. However, our study did not collect data on blood transfusion practices, limiting our ability to confirm this hypothesis. Future research incorporating transfusion data will be essential to validate and further explore this potential explanation.

In our study, hypoglycemia significantly increased the instantaneous risk of death by more than twofold compared to children with normal blood glucose levels. Severe infections such as pneumonia represent hypermetabolic states characterized by elevated glucose demands. While energy requirements rise, intake is often reduced due to impaired consciousness, anorexia, and persistent vomiting. These conditions can lead to hypoglycemia, which may result in cellular dysfunction, particularly cerebral, manifesting as convulsions and further compromising survival. Hypoglycemia may also precipitate life-threatening multiorgan failure. Co-infection with malaria, which is particularly hypoglycemic due to increased glucose consumption by Plasmodium parasites, may exacerbate this condition. This creates a vicious cycle in which hypoglycemia worsens the effects of infection, and the infection, in turn, deepens hypoglycemia, amplifying the risk of death. Monitoring blood glucose levels in children hospitalized for severe acute infections, including severe pneumonia, is therefore critical. A comprehensive management strategy combining appropriate nutritional support and rigorous clinical monitoring is essential to reduce mortality in this high-risk population.

In our study, three main antibiotics (ceftriaxone, gentamicin, ampicillin) were used alone or in combination. The combination of ceftriaxone and gentamicin was the most commonly prescribed antibiotic regimen. However, this widespread use of ceftriaxone is not in line with WHO recommendations for the management of severe pneumonia, which advocate dual antibiotic therapy with ampicillin and gentamicin as first-line treatment. According to these guidelines, ceftriaxone should be reserved for second-line use, either after treatment failure or in cases where HIV infection is suspected or confirmed [7]. Ampicillin and ceftriaxone alone or in combination with gentamicin significantly reduced the immediate risk of death. This protective effect was more pronounced with the combination of ceftriaxone and gentamicin [aSHR = 0,45; IC95%: (0,30-0,69)]. This result is consistent with data from previous studies indicating that third-generation cephalosporin-gentamicin combinations are associated with improved survival in cases of severe pneumonia in children [28].

Our study has some limitations. First, certain characteristics potentially associated with pneumonia-related mortality were not included in our analysis, such as parental factors (e.g., maternal education level, place of residence, household wealth index), prior medical history [29,30], delay in care-seeking [31], quality of care provided [32,33], antibiotic changes during hospitalization, the causative agent [11], and radiographic findings [34,35]. Second, the diagnosis of severe pneumonia was based on WHO criteria [7], which lack specificity. Misclassification bias may have occurred, particularly in relation to malaria—a highly endemic disease in the region—whose severe clinical signs often overlap with those of severe pneumonia. Third, our study was conducted in a secondary-level referral hospital. As such, the findings may not be generalizable to primary-level facilities or to the broader population. Fourth, mortality could not be assessed in children who were referred to other facilities, which may have led to an underestimation of pneumonia-related mortality. Despite these limitations, our study fills a critical data gap on in-hospital mortality related to severe pneumonia among children in our context. Major strengths include the large sample size and the extended data collection period spanning nearly 30 months.

## Conclusion

Although substantial progress has been made in controlling childhood infectious diseases in Burkina Faso, severe pneumonia remains a major concern due to its high hospitalization rate and elevated mortality. In our study, several factors were significantly associated with an increased risk of death, including younger age, hospitalization during the rainy season, hypoxemia, convulsions, stridor, and hypoglycemia. Antibiotic therapy was found to have a protective effect. Additionally, anemia emerged as a protective factor, which may reflect the beneficial impact of blood transfusion on the survival of critically ill children but requires further research to explain. These findings underscore the importance of effective triage at admission and the implementation of targeted management strategies that prioritize care for the most vulnerable children.

## Data Availability

The data underlying this study are owned by the Ministry of Health of Burkina Faso and cannot be made publicly available due to institutional restrictions. Data are stored and managed on the STELab platform (Système de Transport des Échantillons de Laboratoire) and are available from the Ministry of Health upon reasonable, motivated request. Interested researchers may submit a data access request through the corresponding author, who will liaise with the Ministry of Health of Burkina Faso to facilitate access via the STELab system.

## Acknowledges

The authors express their sincere acknowledges to the Ministry of Health of Burkina Faso for granting permission to use data from the national health information system. They also thank the U.S. Centers for Disease Control and Prevention, Atlanta, for the technical and financial support provided to the Ministry of Health in implementing severe pneumonia surveillance in several hospitals across the country. The authors are particularly grateful to the staff of the pediatric department of the Banfora Regional Hospital, especially the pneumonia surveillance focal points — Roseline Ilboudo, Noaga Konkobo, Arnaud Sanou, Abdou Razak Ouédraogo, and Boukaré Sawadogo — for their valuable contribution to patient enrolment and case report form completion.

## Author Contributions

| Contributor Role | Authors |
| --- | --- |
| Conceptualization | Kiebré Pegd-Wendé Blaise, Desiré Lucien Dahourou |
| Methodology | Kiebré Pegd-Wendé Blaise, Desiré Lucien Dahourou |
| Data Curation | Bountogo Mamadou, Sawadogo Guetwendé, Zoma Robert |
| Formal Analysis | Kiebré Pegd-Wendé Blaise ; Desiré Lucien Dahourou |
| Supervision | Meda Nicolas |
| Validation | Meda Nicolas |
| Visualization | Kiebré Pegd-Wendé Blaise, Desiré Lucien Dahourou |
| Writing – Original Draft Preparation | Kiebré Pegd-Wendé Blaise, Desiré Lucien Dahourou |
| Writing – Review & Editing | Bountogo Mamadou, Dah Ter Tiero Elias, Tall Haoua, Boere Djibril, Ouedraogo Achille, Konaté Zanga Youssouf, Sawadogo Guetwendé, Zoma Robert, Meda Nicolas |

All authors have read and agreed to the published version of the manuscript.

